# 3D Capsule Networks for Brain Image Segmentation

**DOI:** 10.1101/2022.01.18.22269482

**Authors:** Arman Avesta, Yongfeng Hui, Mariam Aboian, James Duncan, Harlan M. Krumholz, Sanjay Aneja

## Abstract

**Background and Purpose:** Current auto-segmentation models of brain structures, UNets and nnUNets, have limitations, including the inability to segment images that are not represented during training and lack of computational efficiency. 3D capsule networks (CapsNets) have the potential to address these limitations.

**Methods:** We used 3430 brain MRIs, acquired in a multi-institutional study, to train and validate our models. We compared our CapsNet with standard alternatives, UNets and nnUNets, based on segmentation efficacy (Dice scores), segmentation performance when the image is not well-represented in the training data, performance when the training data are limited, and computational efficiency including required memory and computational speed.

**Results:** The CapsNet segmented the third ventricle, thalamus, and hippocampus with Dice scores of 95%, 94%, and 92%, respectively, which were within 1% of the Dice scores of UNets and nnUNets. The CapsNet significantly outperformed UNets in segmenting images that are not well-represented in the training data, with Dice scores 30% higher. The computational memory required for the CapsNet is less than a tenth of the memory required for UNets or nnUNets. The CapsNet is also more than 25% faster to train compared with UNet and nnUNet.

**Conclusion:** We developed and validated a CapsNet that is effective in segmenting brain images, can segment images that are not well-represented in the training data, and are computationally efficient compared with alternatives.

## Introduction

Neuroanatomical image segmentation is an important component in the management of various neurological disorders.^1–3^ Accurate segmentation of anatomical structures on brain magnetic resonance images (MRIs) is an essential step in a variety of neurosurgical and radiation therapy procedures.^1,3–6^ Manual segmentation is time-consuming and is prone to intra- and inter-observer variability.^7,8^ With the advent of deep learning to automate various image analysis tasks,^9,10^ there has been increasing enthusiasm to use deep learning for brain image auto-segmentation.^11–14^

UNets are among the most popular and successful deep learning auto-segmentation algorithms.^11,15,16^ Despite the broad success of UNets in segmenting anatomical structures across various imaging modalities, they have well described limitations. UNets perform best on images that closely resemble the images used for training, but underperform on images that contain variant anatomy or pathologies that change the appearance of normal anatomy.^8^ Additionally, UNets have a large number of trainable parameters, hence training and deploying UNets for image segmentation often requires substantial computational resources that may not be scalable in all clinical settings.^15^ There is a need for fast, computationally efficient segmentation algorithms that can segment images not represented in the training data with high fidelity.

Capsule networks (CapsNets) represent an alternative auto-segmentation method that can potentially overcome the limitations of UNets.^17–19^ CapsNets can encode and manipulate spatial information such as location, rotation, and size about structures within an image, and use this spatial information to produce accurate segmentations. Encoding spatial information allows CapsNets to generalize well on images that are not well-represented in the data used to train the algorithm.^18,19^ Moreover, CapsNets use a smarter paradigm for information encoding which relies on fewer parameters leading to increased computational efficiency.^17–19^

Capsule networks have shown promise on some biomedical imaging tasks,^19^ but have yet to be fully explored for segmenting anatomical structures on brain MRIs. In this study, we explore the utility of CapsNets for segmenting anatomical structures on brain MRIs using a multi-institutional dataset of more than 3,000 brain MRIs. We compare the segmentation efficacy and computational efficiency of CapsNets with popular UNet-based models.

## Methods

### Dataset

The dataset for this study included 3,430 T1-weighted brain MRI images, belonging to 841 patients from 19 institutions enrolled in Alzheimer’s Disease Neuroimaging Initiative (ADNI) study.^20^ The inclusion criteria of ADNI have been previously described.^21^ On average, each patient underwent four MRI acquisitions. Details of MRI acquisition parameters are provided in Appendix 1.^20^ We randomly split the patients into training (3,199 MRI, to 93% of data), validation (117 MRI volumes, 3.5% of data), and test (114 MRI volumes, 3.5% of data) sets. Data was divided at the patient level to assure that all images belonging to a patient were assigned either the training, validation, or test set. Patient demographics are provided in Table 1. This study was approved by the Institutional Review Board of Yale School of Medicine (IRB number 2000027592).

**Table 1:** Study participants. tabulated by the training, validation, and test sets.

| Data Partitions | Number of<br>MRI volumes | Number of<br>patients | Age<br>mean ± SD | Gender<sup>†</sup> | Diagnosis<sup>††</sup> |
| --- | --- | --- | --- | --- | --- |
| Training set | 3199 | 841 | 76 ± 7 | 42% F, 58% M | 29% CN, 54% MCI, 17% AD |
| Validation set | 117 | 30 | 75 ± 6 | 30% F, 70% M | 21% CN, 59% MCI, 20% AD |
| Test set | 114 | 30 | 77 ± 7 | 33% F, 67% M | 27% CN, 47% MCI, 26% AD |

<sup>†</sup> F: female; M: male.
<sup>††</sup> CN: cognitively normal; MCI: mild cognitive impairment; AD: Alzheimer’s disease.

### Anatomic Segmentations

We trained our models to segment three anatomical structures of the brain: third ventricle, thalamus, and hippocampus. These structures were chosen to represent structures with varying degrees of segmentation difficulty. Preliminary ground-truth segmentations were initially generated using FreeSurfer,^22–24^ and then manually corrected by a board-eligible radiologist with nine years of experience in neuroimaging research.

### Image Pre-Processing

MRI preprocessing included correction for intensity inhomogeneities, including B1-field variations.^25,26^ The 3D brain volume was cropped after removing the skull, face, and neck tissues ^27^ To overcome memory limitations, segmentations were done on 64×64×64-voxel patches of the MRI volume that contained the segmentation target. The patch was automatically placed over the expected location of the segmentation target using pre-defined coordinates referenced from the center of the image. The coordinates of each patch were computed during training and were fixed during testing, without any manual input and without using the ground-truth segmentations. Details of pre-processing are provided in Appendix 2.

### Capsule Networks

CapsNets are composed of three main components: 1) capsules that each encode a structure together with the *pose* of that structure: the pose is an n-dimensional vector that learns to encode orientation, size, curvature, location, and other spatial information about the structure; 2) a supervised learning paradigm that learns how to transform the poses of the parts (e.g. head and tail of hippocampus) to the pose of the whole (e.g. the entire hippocampus); and 3) a clustering paradigm that detects a whole if the poses of all parts transform into matching poses of the whole. Further details regarding differences between CapsNets and other deep learning models are provided in Appendix 3.

2D CapsNets were previously introduced by LaLonde et al to segment one slice of the image at a time.^19^ We developed 3D CapsNets for volumetric segmentation of a 3D volume, with the architecture shown in Figure 1.A. The first layer, Conv1, performs 16 convolutions (5×5×5) on the input volume to generate 16 feature volumes, which are reshaped into 16D vectors at each voxel. The 16D vector at each voxel is reshaped into a pose that learns to encode spatial information at that voxel. The next layer, PrimaryCaps2, has two capsule channels that learn two 16D-to-16D convolutional transforms (5×5×5) from the poses of the previous-layer parts to the poses of the next-layer wholes. Likewise, all capsule layers (green layers in Figure 1.A) learn m-to-n-dimensional transforms from the poses of parts to the poses of wholes. Our CapsNet has downsampling and upsampling limbs. The downsampling limb learns *what* structure is present at each voxel, and the skip connections from downsampling to upsampling limbs preserve *where* each structure is on the image. Downsampling is done using 5×5×5 convolutional transforms with stride = 2. Layers in the deeper parts of CapsNet contain more capsule channels (up to 8) and poses with more components (up to 64) to be able to encode more complex structures, since each capsule in the deeper parts of the model should be able to detect complex concepts in the entire image. Upsampling is done using 4×4×4 transposed convolutional transforms with stride = 2 (turquoise layers in Figure 1A). The final layer, FinalCaps13, contains one capsule channel that learns to activate capsules within the segmentation target and deactivate them outside the target. Appendix 4 explains the options that we explored for developing our 3D CapsNets and how we chose the best design options. Appendix 5 explains how the final layer activations were converted into segmentations. Details about how the model finds agreeing poses of parts that vote for the pose of the whole are provided in Appendix 6.

**Figure 1:**
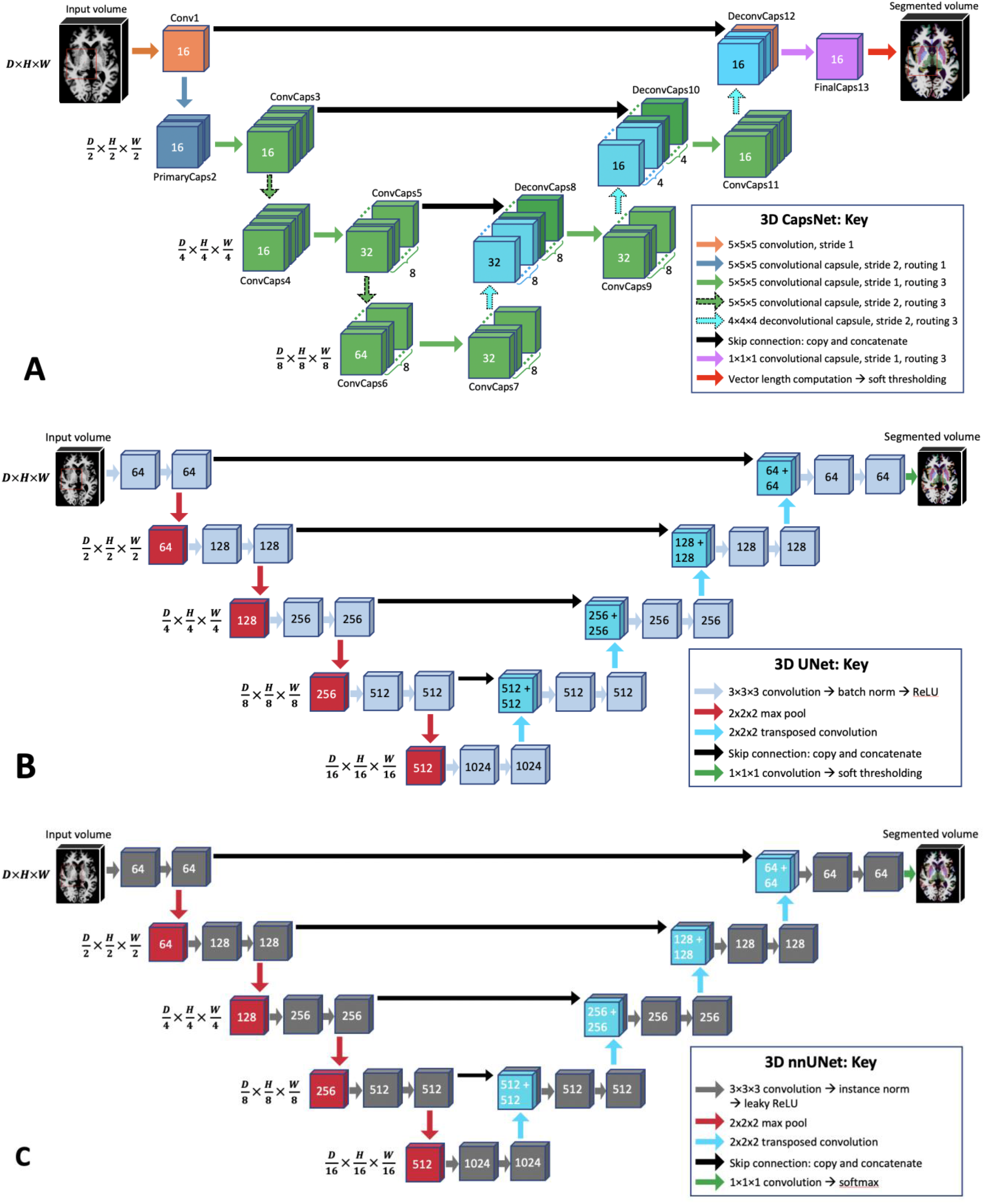
CapsNet (A), UNet (B), and self-configured nnUNet (C) architectures. All models process 3D images in all layers, with dimensions shown on the left side. *D, H*, and *W* respectively represent the depth, height, and width of the image in each layer. In (A), the number over the Conv1 layer represents the number of channels. The numbers over the capsule layers (ConvCaps, DeconvCaps, and FinalCaps) represent the number of pose components. The stacked layers represent capsule channels. In (B) and (C), the numbers over each layer represent the number of channels. In UNet and nnUNet, the convolutions have stride=1 and the transposed convolutions have stride = 2. Please note that the numbers over capsule layers show the number of pose components, while the numbers over non-capsule layers show the number of channels.

### Comparisons: UNets and nnUNets

Optimized 3D UNets and nnUNets were also trained on the same training data, ^11–13,28^ and their segmentation efficacy and computational efficiency were compared with our CapsNet using the same test data. UNets and nnUNets have shown strong auto-segmentation performance across a variety of different imaging modalities and anatomic structures and are among the most commonly used segmentation algorithms in biomedical imaging.^11– 13,15,29,30^ Figure 1.B shows the architecture of our UNet. The input image undergoes 64 convolutions (3×3×3) to generate 64 feature maps. These maps then undergo batch normalization and ReLU activation. Similar operations are carried out again, followed by downampling using max-pooling (2×2×2). The downsampling and upsampling limbs each include four units. Upsampling is done using 2×2×2 transposed convolutions with stride = 2. The final layer carries out a 1×1×1 convolution to aggregate all 64 channels, followed by soft thresholding using the sigmoid function. The model learns to output a number close to 1 for each voxel inside the segmentation target, and a number close to 0 for each voxel outside the target. We also trained self-configuring nnUNets that automatically learn the best architecture as well as the optimal training hyperparameters.^16^ Figure 1.C shows the architecture of the nnUNet resulting from the self-configuring nnUNet paradigm.

### Model Training

The CapsNet and UNet models were trained for 50 epochs using Dice loss and the Adam optimizer.^31^ Initial learning rate was set at 0.002. We used dynamic paradigms for learning rate scheduling, with a minimal learning rate of 0.0001. The hyperparameters for our UNet were chosen based on the best-performing model over the validation set. The hyperparameters for nnUNet were self-configured by the model.^16^ The training hyperparameters for CapsNet and UNet are detailed in Appendix 6.

### Model Performance

The segmentation efficacy of the three models was measured using Dice scores. To compare the performance of each segmentation model when training data is limited, we also trained the models using subsets of the training data with 600, 240, 120, and 60 MRIs. We then compared the segmentation efficacy of the models using the test set.

The relative computational efficiency of the models was measured by 1) the computational memory required to run the model (in megabytes), 2) the computational time required for training each model, and 3) the time that each model takes to segment one MRI volume.

### Out-of-Distribution Testing

To evaluate the performance of CapsNet and UNet models on the images that were not represented during training, we trained the models using images of the right hemisphere of the brain that only contained the right thalamus and right hippocampus. Then, we evaluated the segmentation efficacy of the trained models on the images of the left hemisphere of the brain that contained the contralateral left thalamus and left hippocampus. Because the left-hemisphere images in the test set are not represented in the right-hemisphere images in the training set, this experiment evaluates the out-of-distribution performance of the models. We intentionally did not use any data augmentation during training to assess out-of-distribution performance of the models. Given that nnUNet paradigm requires data augmentation, nnUNet was not included in this experiment.

### Implementation

Image pre-processing was done using Python (version 3.9) and FreeSurfer (version 7). PyTorch (version 1.11) was used for model development and testing. Training and testing of the models were run on GPU-equipped servers (4 vCPUs, 61 GB RAM, 12 GB NVIDIA GK210 GPU with Tesla K80 Accelerators). Code used to train and test our models, our pre-trained models, and a sample MRI are available on our lab’s GitHub page: www.github.com/Aneja-Lab-Yale/Aneja-Lab-Public-CapsNet.

## Results

All three segmentation models showed high performance across all three neuroanatomical structures with Dice scores above 90% (Table 2). Performance was highest for the 3^rd^ ventricle (95%-96%) followed by the thalamus (94%-95%) and hippocampus (92%-93%). Dice scores between the CapsNet and UNet-based models were within 1% for all neuroanatomical structures.

**Table 2:** Comparing the segmentation efficacy of CapsNets, UNets, and nnUNets in segmenting brain structures that were represented in the training data. The segmentation accuracy was quantified using Dice scores on the test (114 brain MRIs). The 3rd ventricle, thalamus, and hippocampus respectively represent easy, medium, and difficult structures to segment.

| Brain structure | CapsNet Dice (95% CI) | UNet Dice (95% CI) | nnUNet Dice (95% CI) |
| --- | --- | --- | --- |
| 3rd ventricle | 95%<br>(94 to 96) | 96 %<br>(95 to 97) | 96%<br>(95 to 97) |
| Thalamus | 94%<br>(93 to 95) | 95 %<br>(94 to 96) | 94%<br>(92 to 96) |
| Hippocampus | 92 %<br>(91 to 93) | 93 %<br>(92 to 94) | 92%<br>(91 to 93) |

Segmentation performance for each model remained high across training datasets of varying sizes (Figure 4). When training on 120 brain MRIs, all three models maintained their segmentation accuracy within 1% when compared to models trained on 3,199 brain MRIs. Segmentation performance did decrease for all three models when trained on 60 brain MRIs (83% CapsNet, 84% UNet, 88% nnUNet)

**Figure 2:**
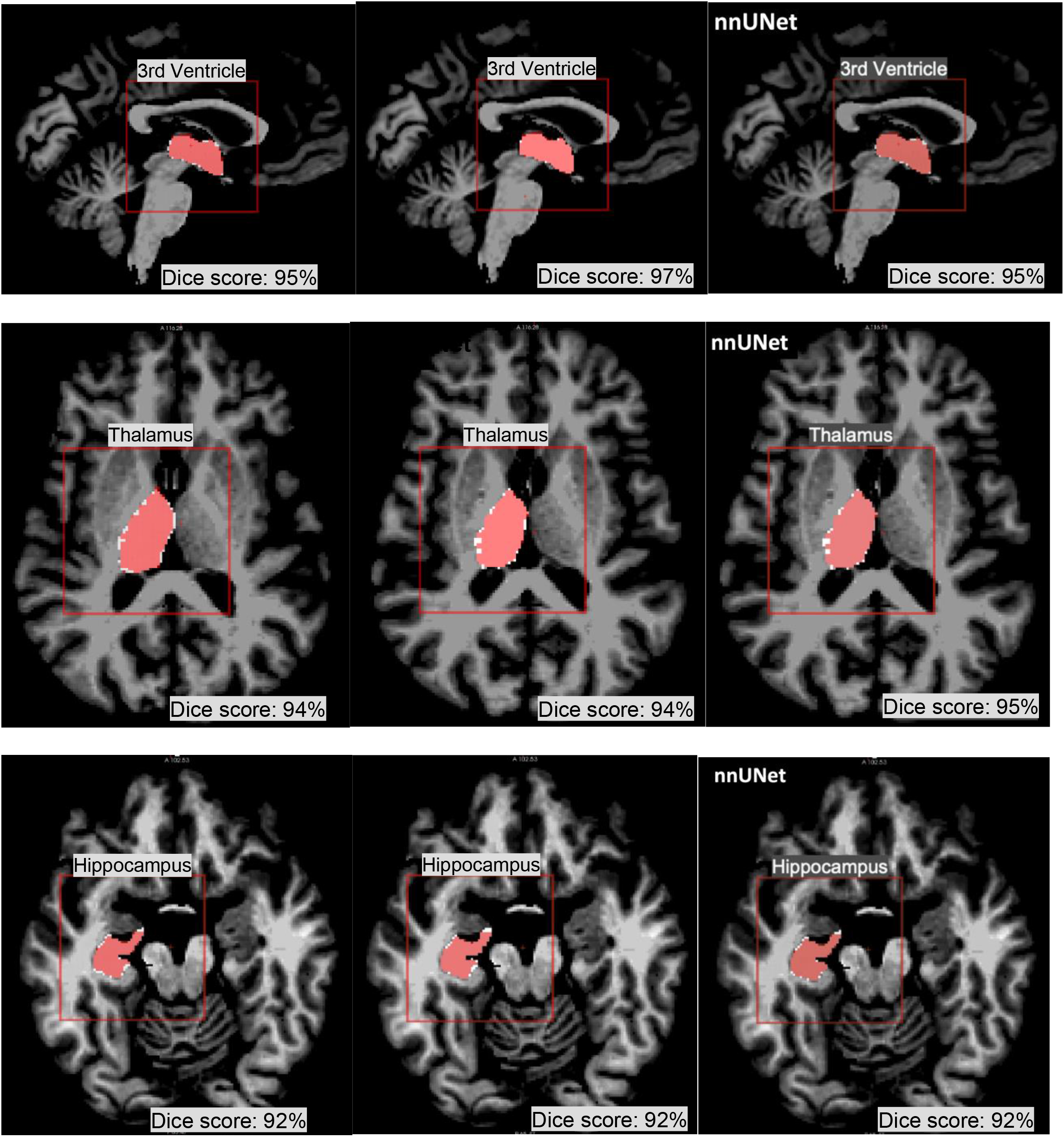
CapsNet, UNet, and nnUNet segmentation of brain structures that *were represented* in the training data. Segmentations for three structures are shown: 3rd ventricle, thalamus, and hippocampus. Target segmentations and model predictions are respectively shown in red and white. Dice scores are provided for the entire volume of the segmented structure *in this patient* (who was randomly chosen from the test set).

**Figure 3:**
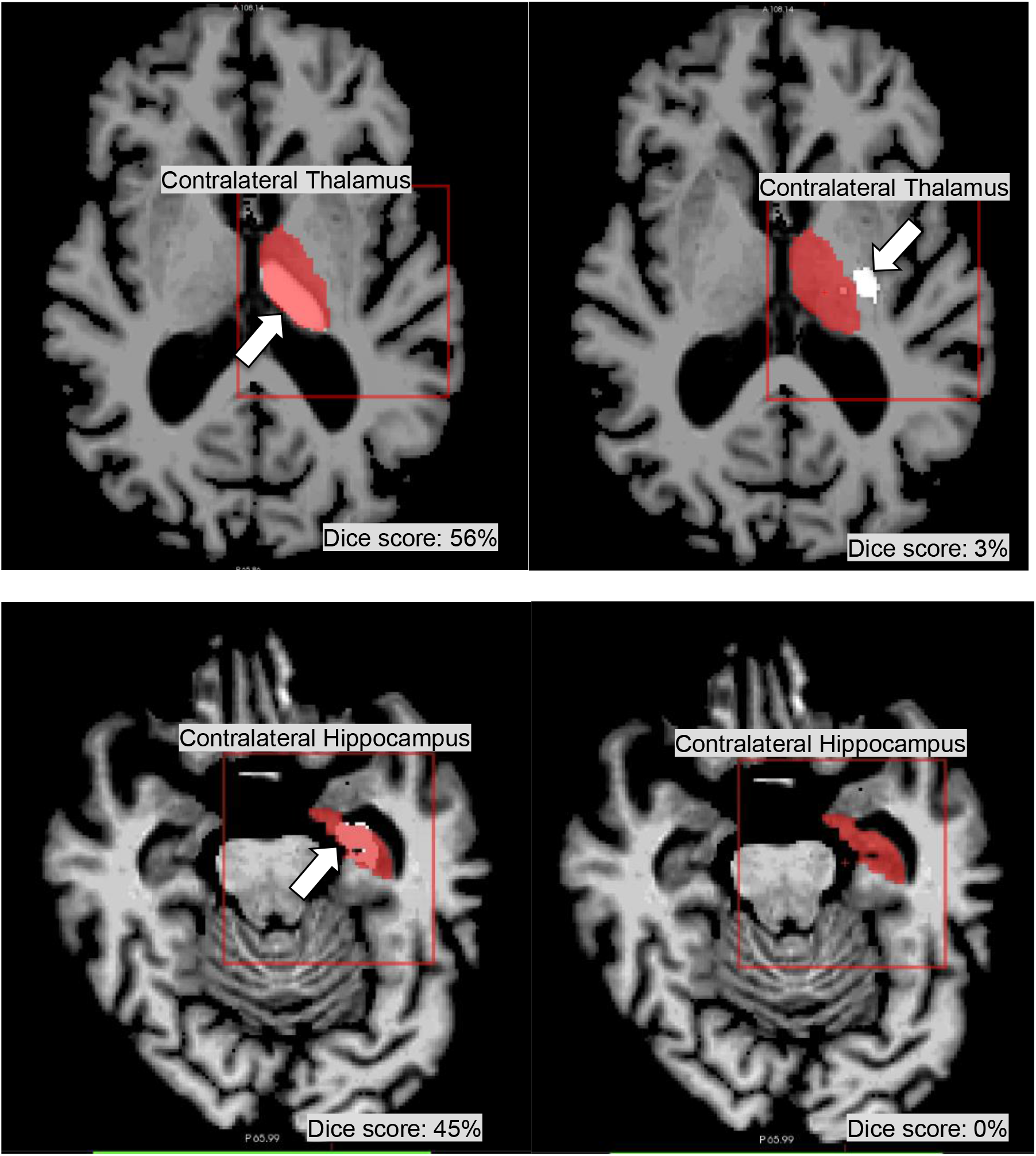
CapsNets outperforms UNets in segmenting images that were not represented in the training data. Both models were trained to segment right brain structures, and were tested to segment contralateral left brain structures. Target segmentations and model predictions are respectively shown in red and white. Dice scores are provided for the entire volume of the segmented structure *in this patient*. The CapsNet partially segmented the contralateral thalamus and hippocampus, but the UNet poorly segmented the thalamus and entirely missed the hippocampus.

**Figure 4:**
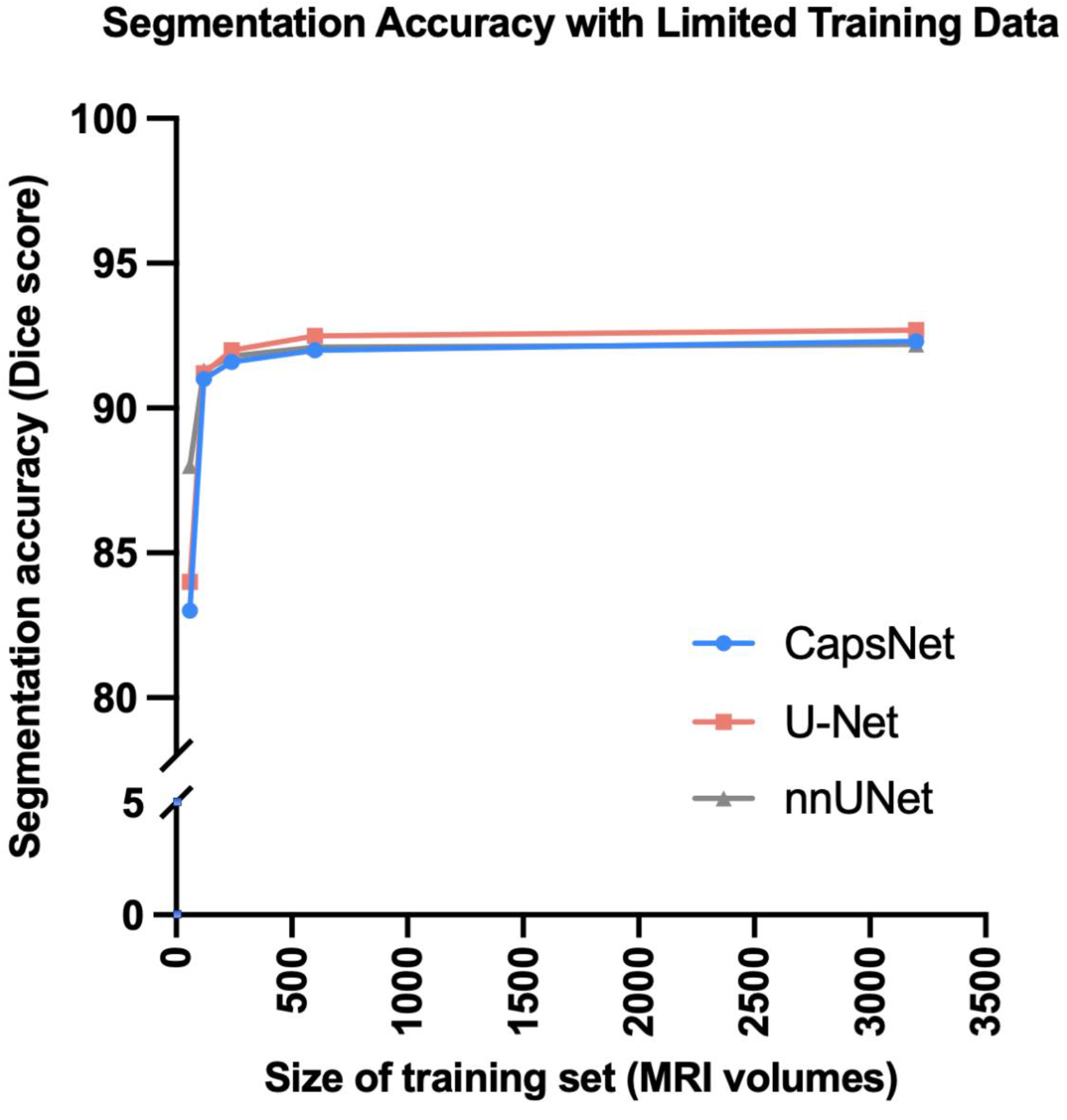
Comparing CapsNets, UNets, and nnUNets when training data is limited. When the size of the training set was decreased from 3199 to 120 brain MRIs, hippocampus segmentation accuracy (measured by Dice score) of all three models did not decrease more than 1%. Further decrease in the size of the training set down to 60 MRIs led to worsened segmentation accuracy.

Although both CapsNet and UNet had difficulty segmenting contralateral structures, the CapsNet significantly outperformed the UNet (Thalamus P-value < 0.001, Hippocampus P-value < 0.001) (Table 3). CapsNet models frequently identified the contralateral structure of interest but underestimated the size of the segmentation, resulting in Dice scores between 40% and 60%. In contrast, the UNet models frequently failed to identify the contralateral structure of interest, resulting in Dice scores lower than 20% (Figure 3).

**Table 3:** Comparing the efficacy of CapsNets and UNets in segmenting images that were not represented in the training data. Both models were trained to segment the right thalamus and hippocampus. Then, they were tested on segmenting the contralateral left thalamus and hippocampus.

| Brain structure | CapsNet Dice (95% CI) | UNet Dice (95% CI) | CapsNet vs UNet P-value<sup>†</sup> |
| --- | --- | --- | --- |
| Thalamus | 52%<br>(46 to 58) | 16%<br>(11 to 21) | < 0.01 |
| Hippocampus | 43%<br>(38 to 48) | 10%<br>(6 to 14) | < 0.01 |

<sup>†</sup> Paired-samples t-test, degrees of freedom = 114 - 1 = 113

The CapsNet was more computationally efficient compared to UNet-based models (Figure 5). The CapsNet required 228 MBs, compared to 1,364 MBs for UNet and 1,410 MBs for nnUNet. The CapsNet trained 25% faster than the UNet (1.5s v 2s per sample) and 100% faster than the nnUNet (1.5s vs 3s per sample). When we compared the deployment times of the fully-trained models, CapsNet and UNet could segment images equally fast (0.9s per sample) which was slightly faster than the nnUNet (1.1s per sample).

**Figure 5:**
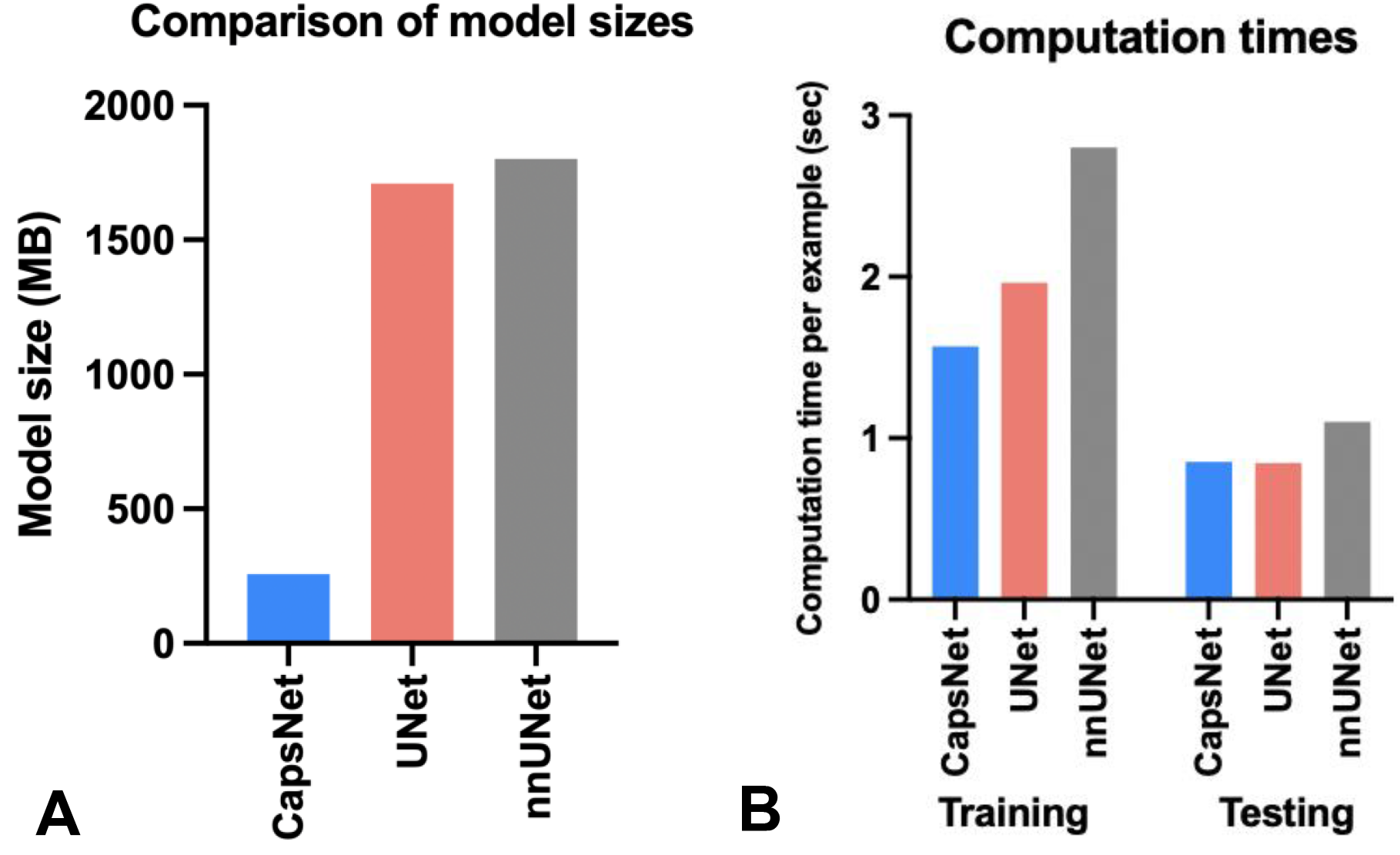
Comparing the computational efficiency between CapsNets, UNets, and nnUNets, in terms of memory requirements (A) and computational speed (B). The bars in (A) represent the computational memory required to accommodate the total size of each model, including the parameters plus the cumulative size of the forward- and backward-pass feature volumes. CapsNet trains faster (B), given that its trainable parameters are one order of magnitude fewer than UNets or nnUNets. The training times represent the time that each model took to converge for segmenting hippocampus, divided by the number of training examples and the training epochs (to make training times comparable with test times). The test times represent how fast a fully-trained model can segment a brain image.

## Discussion

Neuroanatomic segmentation of brain structures is an essential component in the treatment of various neurologic disorders. Deep-learning-based auto-segmentation methods have shown the ability to segment brain images with high fidelity, which was previously a time-intensive task.^13,14^ In this study, we compared the segmentation efficacy and computational efficiency of CapsNets with UNet-based auto-segmentation models. We found CapsNets to be reliable and computationally efficient, achieving segmentation accuracy comparable to commonly-used UNet-based models. Moreover, we found CapsNets to have higher segmentation performance on out-of-distribution data, suggesting an ability to generalize beyond their training data.

Our results corroborate previous studies demonstrating the ability of deep learning models to reliably segment anatomical structures on diagnostic images.^11,12,14^ UNet-based models have been shown to effectively segment normal anatomy across a variety of different imaging modalities including CT, MRI, and Xray images.^15,29,30,32–34^ Moroever, Isensee et al showed the ability of nnUNets to generate reliable segmentations across 23 biomedical image segmentation tasks with automated hyperparameter optimization. We have extended prior work by demonstrating similar segmentation efficacy between CapsNets and UNet-based models with CapsNets being notably more computationally efficient. Our CapsNets require less than 10% the amount of memory required by UNet-based methods and trains 25% faster.

Our findings are consistent with prior studies demonstrating the efficacy of CapsNets for image segmentation.^19,35^ LaLonde et al previously demonstrated that 2D CapsNets can effectively segment lung tissues on CT images and muscle and fat tissues on thigh MR images. Their group similarly found that CapsNets can segment images with performance rivaling UNet-based models while requiring with less than 10% of the memory required by UNet-based models. Our study builds upon prior studies by showing the efficacy of CapsNets for segmenting neuroanatomical substructures on brain MRIs. Additionally, compared to prior work, we have implemented 3D CapsNet architecture, which has not been previously described in the literature.

Previous studies have suggested that CapsNets are able to generalize beyond their training data.^18,19^ Hinton et al demonstrated that CapsNets can learn spatial information about the objects in the image, and can then generalize this information beyond what is present in the training data, which gives CapsNet out-of-distribution generalization capability.^18^ The ability to segment out-of-distribution images was also shown by LaLonde et al for their 2D CapsNet model that segments images.^19^ We build upon previous studies by demonstrating out-of-distribution generalizability of 3D CapsNets for segmenting medical images.

Although we found CapsNets to be effective in biomedical image segmentation, previous studies on biomedical imaging have shown mixed results.^35^ Survarachakan et al previously found 2D CapsNets to be effective for segmenting heart structures, but ineffective for segmenting hippocampus on brain images.^35^ Our more favorable results in segmenting hippocampus are likely because of the 3D structure of our CapsNet, which can use the contextual information in the volume of the image rather than just a slice of the image, to better segment the complex shape of the hippocampus.

Our study has several limitations which should be noted. Our models were only tested on three brain structures that are commonly segmented on brain MRIs, meaning that our findings may not generalize across other imaging modalities and anatomic structures. Nevertheless, our findings show the efficacy of CapsNets on brain structures with different levels of segmentation difficulty, suggesting potential utility for a variety of scenarios. Computational efficiency across models was measured using the same computing resources and GPU memory, and our findings may not translate to different computational settings. Future studies can further explore the relative computational efficiency of CapsNets compared to other auto-segmentation models across different computing environments. Lastly, we only compared the efficacy of CapsNets withUNet-based models. While there are multiple other auto-segmentation models, UNet-based models are currently viewed as the most successful deep-learning models for segmenting biomedical images. Further studies comparing the CapsNet to other deep-learning models are an area of future research.

## Conclusion

In this study, we showed that 3D CapsNets can accurately segment neuroanatomical structures on brain MR images with segmentation accuracy similar to UNet-based models. We also showed that CapsNets outperform UNet-based models in segmenting out-of-distribution data. CapsNets are also more computationally efficient compared to UNet-based models, since they train faster and require less computation memory.

## Data Availability

All data produced are available online at: 
adni.loni.usc.edu

## Abbreviations

ADNI: Alzheimer’s disease neuroimaging initiative
CapsNet: capsule network
CPU: central processing unit
CT: computed tomography
GB: giga-byte
GPU: graphics processing unit
MRI: magnetic resonance imaging

## Acknowledgements

Arman Avesta is a PhD Student in the Investigative Medicine Program at Yale which is supported by CTSA Grant Number UL1 TR001863 from the National Center for Advancing Translational Science, a component of the National Institutes of Health (NIH). The contents of this article are solely the responsibility of the authors and do not necessarily represent the official view of NIH. This work was supported by the Radiological Society of North America’s Fellow Research Grant Number RF2212RO. The data used in this article were obtained from the Alzheimer’s Disease Neuroimaging Initiative (ADNI) database (adni.loni.usc.edu). The investigators within the ADNI contributed to the design and implementation of ADNI but did not participate in the analysis or writing of this article.

## Disclosures

### Arman Avesta

Arman Avesta is on the trainee editorial board of *Radiology: Artificial Intelligence*. Journal policy recused the author from having any role in the peer review of this manuscript.

### Research Funding

CTSA UL1 TR001863 from the National Center for Advancing Translational Science.

### Yongfeng Hui

The publication was written prior to Yongfeng Hui joining Amazon.

### Mariam Aboian

**Research funding:** KL2 TR001862 from the National Center for Advancing Translational Science and NIH roadmap for Medical Research. The contents of this article are solely the responsibility of the authors and do not necessarily represent the official view of NIH.

### James S. Duncan

**Patents, Royalties, Other Intellectual Property:** Systems, Methods and Apparatuses for Generating Regions of Interest from Voxel Mode Based Thresholds, Publication No: US20190347788A1, application No. 15/978,904. Filed on May 14, 2018, Publication Date: November 14, 2019. Inventors: Van Breugel J, Abajian A, Treli-hard J, Smolka S, Chapiro J, Duncan JS and Lin M. Joint application from Philips, N.V. and Yale University. US Patent 10,832,403 (2020)

### Harlan M. Krumholz

**Employment:** Hugo Health (I), FPrime

**Stock and Other Ownership Interests:** Element Science, Refactor Health, Hugo Health

**Consulting or Advisory Role:** UnitedHealthcare, Aetna

**Research Funding:** Johnson and Johnson

**Expert Testimony:** Siegfried and Jensen Law Firm, Arnold and Porter Law Firm, Martin/Baughman Law Firm

### Sanjay Aneja

**Research Funding:** The MedNet, Inc, American Cancer Society, National Science Foundation, Agency for Healthcare Research and Quality, National Cancer Institute, ASCO, The Patterson Trust

**Patents, Royalties, Other Intellectual Property:** Provisional patent of deep learning optimization algorithm

**Travel, Accommodations, Expenses:** Prophet Consulting (I), Hope Foundation

**Other Relationship:** NRG Oncology Digital Health Working Group, SWOG Digital Engagement Committee, ASCO mCODE Technical Review Group, Associate Editor for *JCO Clinical Cancer Informatics*

## SUPPLEMENTAL MATERIAL

## Appendix 1: MRI acquisition parameters

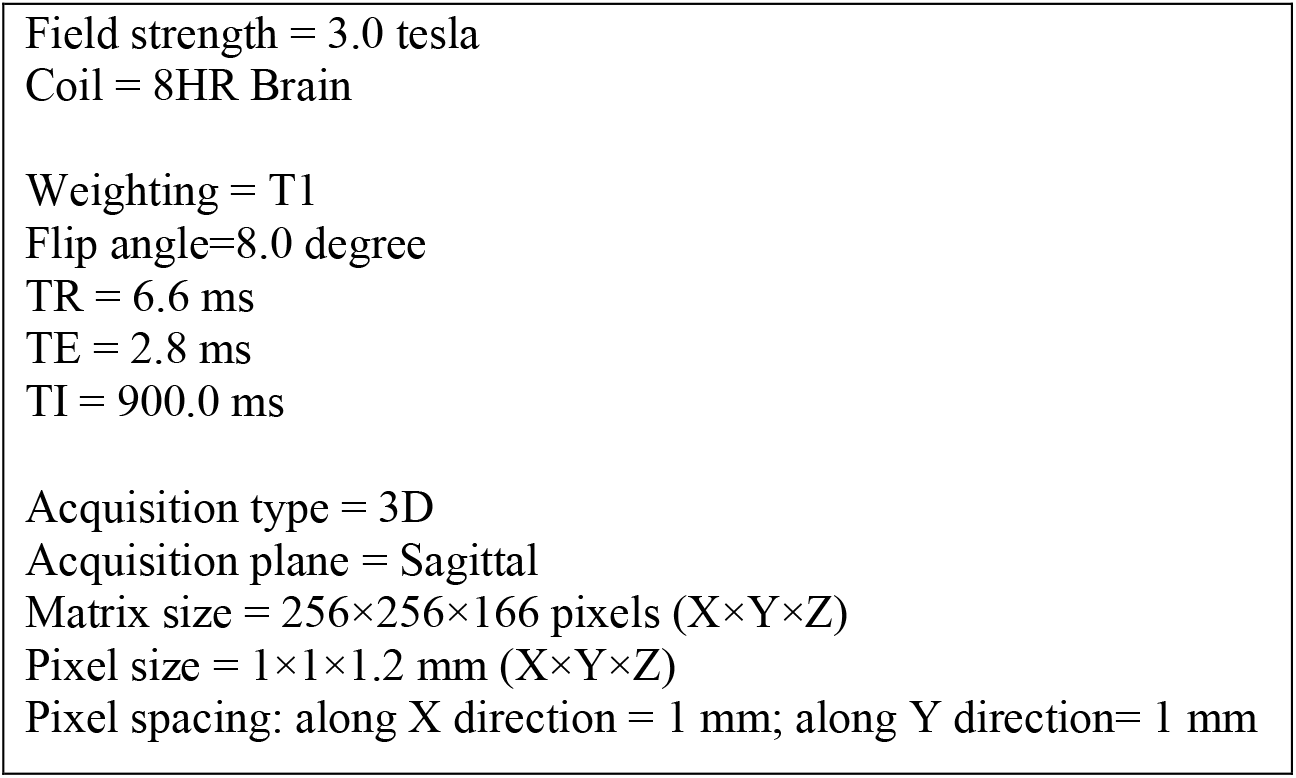

## Appendix 2: Pre-Processing

We corrected for intensity inhomogeneities including B1-field variations. Our pre-processing pipeline first registers the brain image to the MNI305 atlas. Then, pixel intensities are used to roughly segment the white matter. The variations in the pixel intensities in the white matter are then used to estimate the B1 field map Finally, B1 bias field correction is done by dividing the pixel intensities by the estimated bias field.^1^

The next step is the removal of the skull, face, and neck, only leaving the brain. We used a hybrid method of skull stripping that combines a watershed algorithm and a deformable surface model.^2^ This method first roughly segments the white-matter based on pixel intensities. Then, watershed algorithms are used to find the gray-white matter junction and the brain surface. Next, a deformable surface model is used to model the brain surface. The curvature of the brain surface at each point is computed, and these curvatures are used to register the brain surface onto an atlas. The atlas is formed by computing the curvatures of the brain sulci and gyri in several subjects. The reconstructed brain surface, registered to the atlas, is then automatically corrected in case the curvatures in a particular region of the surface do not make sense. The resulting corrected brain surface model is used for skull stripping.^2^

To overcome memory limitations, we cropped 64×64×64-voxel boxes of the MRI volume that contained each segmentation target. The box is automatically placed over the expected location of each segmentation target based on the expected coordinates of the segmentation target. The box is large enough to accommodate inter-subject variability in the expected location of each segmentation target.

**Figure S3:**
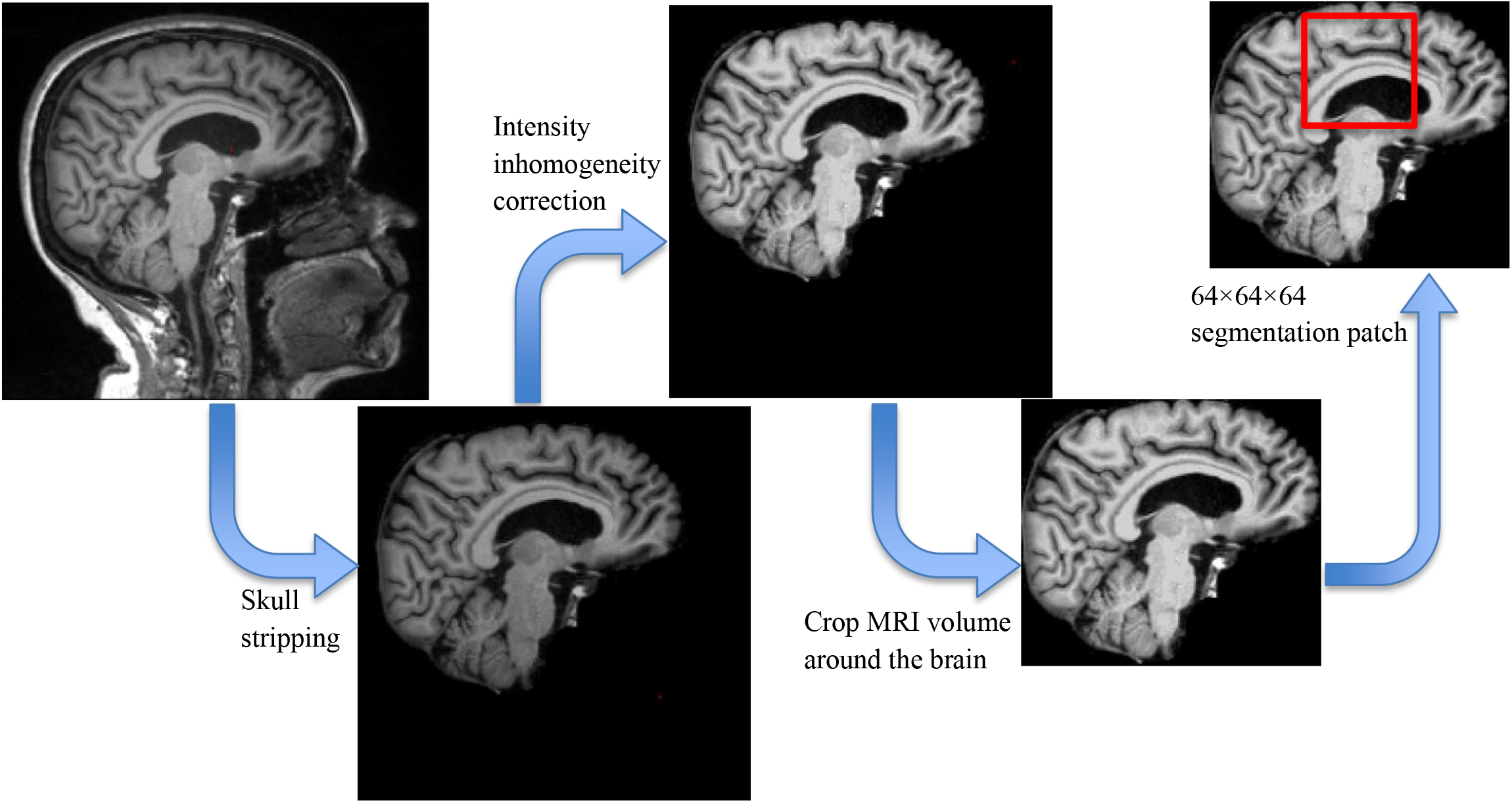
Pre-processing steps.

## Appendix 3: Capsule Networks

Capsule networks (CapsNets) can detect objects when their spatial features change.^3^ This is a fundamental property of CapsNets that enables them to perform well when a test example is not represented in the training data. (A) shows the sagittal T1-weighted brain MRI of a patient with a forward head tilt, and (B) shows the MRI of another patient with a backward head tilt. White arrows (connecting the posterior commissure to the anterior commissure) demonstrate the orientation of the brain. Let’s assume that we have a CapsNet that is trained to segment the entire brain. Let’s also assume that the training set only C contains patients with forward head tilt (like in A). An ideal CapsNet should generalize to segment the brain in patients with a backward tilt (like in B). To achieve this goal, CapsNets encode the spatial features of each structure that they detect. The spatial features of the brain are encoded in a *pose* vector. The pose contains spatial features such as orientation, position, size, curvature, etc. Here, the orientation of the brain (one of the spatial features) is shown by the white arrow. Our goal is to illustrate how CapsNets detect a whole (the brain) when parts (frontal pole, corpus callosum, brainstem, cerebellum, occipital pole, etc.) all vote for the same spatial features of the whole.

CapsNets are composed of three main ingredients: 1) capsules that each encode a structure together with the *pose* of that structure; 2) a supervised learning paradigm that learns the transforms between the poses of the parts (e.g. corpus callosum, brainstem) and the pose of the whole (e.g. the entire brain); and 3) a clustering paradigm that detects a whole if the poses of all parts (after getting transformed) vote for matching poses of the whole. Therefore, any CapsNet needs to: 1) learn the transforms between the poses of parts and wholes; and 2) cluster the votes of the parts to detect wholes.

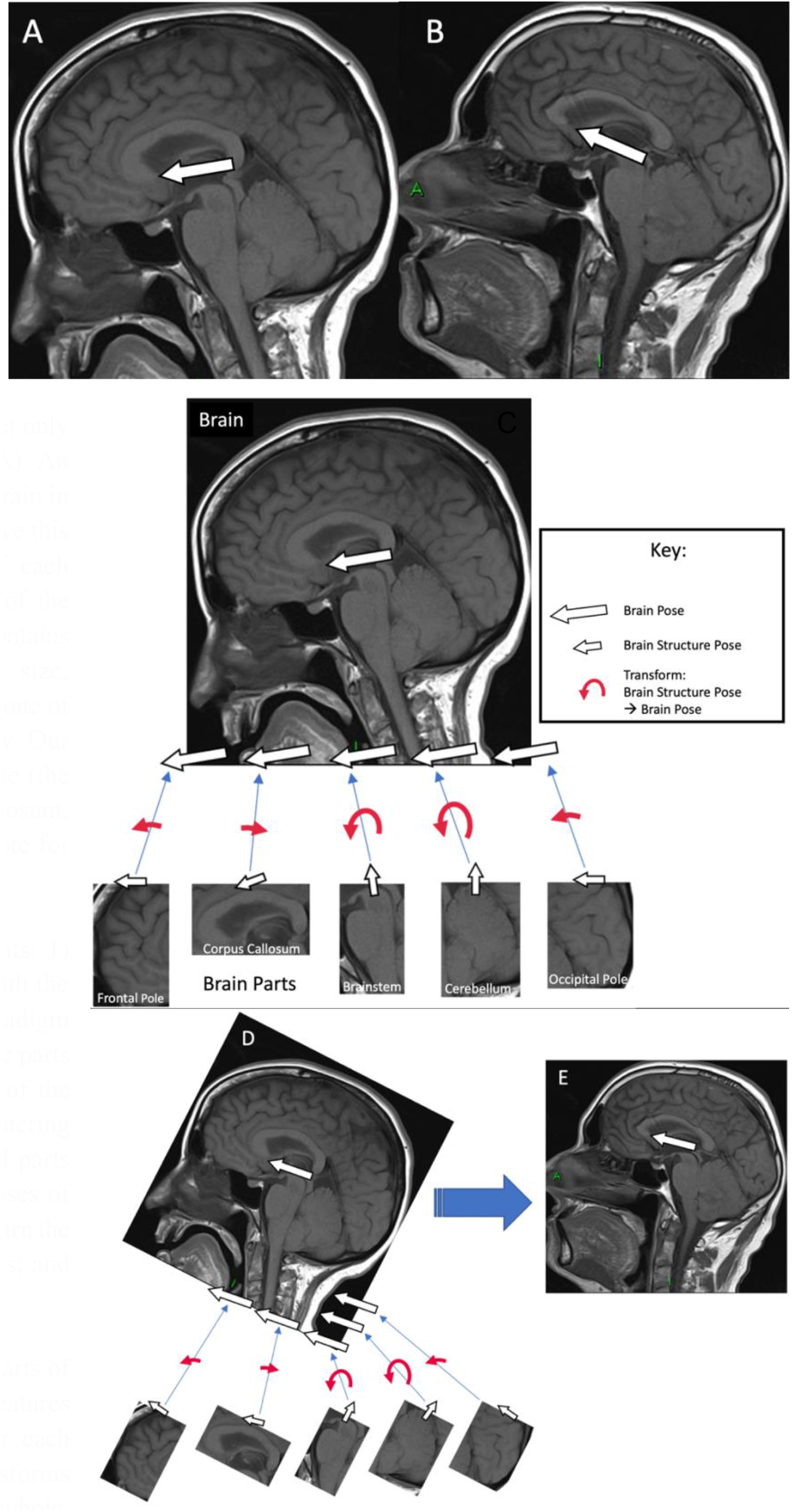

(C) shows a CapsNet that has already detected parts of the brain and has encoded their spatial features (demonstrated by the smaller white arrow over each part). The red curved arrows demonstrate the transforms between the poses of the part and the pose of the whole. After transformation, each part votes for a candidate pose of the whole. If all these votes match, the whole is present. Please note that we are only showing the orientations here for simplicity, but the pose vectors encode more complex spatial features.

In (E), We want the CapsNet to detect the backward-tilted brain while the model is only trained on forward-tilted brain images (such as in C). We can imagine that (E) is just the rotated version of (C), as demonstrated in (D). The parts are all rotated clockwise (compared to the poses of the parts in C). However, the same transforms (red curved arrows) can still transform the poses of the parts into the candidate poses of the whole. The candidate poses of the whole still match, and therefore the whole is detected. This process does not need any data augmentation: an ideal CapsNet can detect objects when they are rotated or have undergone other spatial changes, without the need for any data augmentation. This is because the CapsNet can still use the same transforms between the parts and the wholes (red curved arrows) even though the input image has rotated. Therefore, a change in the poses of the parts will cause an equivalent change in the pose of the whole, while the relationship between the poses of the parts and the whole remains the same. This is a powerful capability that makes CapsNets *equivariant* to the changes in the inputs: spatial change in the inputs will cause an equivalent spatial change in the pose of the detected objects.^3^ Such CapsNets can still detect the changed objects and will encode these changes in the pose of the detected objects. As a result, a CapsNet that is trained on forward-tilted brains (such as in C) can detect backward-tilted brains (such as in E) without the need for any data augmentation.

This approach is fundamentally different from other machine learning methods such as U-Nets (G), which do not have equivariance capabilities. Instead, the max-pooling layers in U-Nets try to kill information about the changes in the inputs to make the model *invariant* to the changed inputs. In essence, CapsNets use equivariance to *encode and model the spatial changes* in the inputs, making CapsNets more efficient in handling variations of the same object.^3^ On the other hand, U-Nets use information killing (in max-pooling layers) to make the model *invariant to the spatial changes* in the inputs. Therefore, U-Nets cannot efficiently detect variations of the same object.

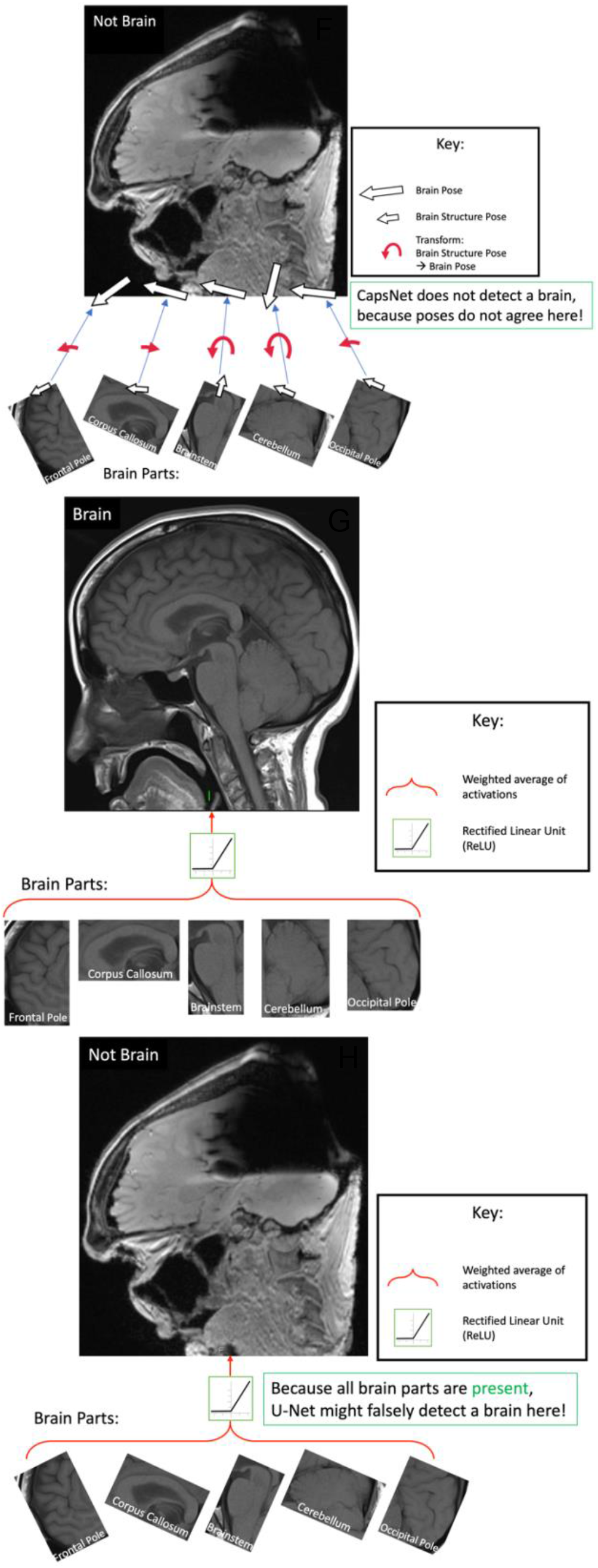

(F) demonstrates why CapsNets are less susceptible to adversarial attacks compared to U-Nets (H). Here, this adversarial image contains all parts of the brain but with orientations that do not make sense, not making a whole. When the poses of the parts are transformed into the candidate poses of the whole (using the same transforms as in C), the candidate poses of the whole do not match. Therefore, the CapsNet would not detect a brain because of the mismatch between the candidate poses of the brain (F). On the other hand, a U-Net that is trained using augmented data may detect the parts. Such a U-Net has no mechanism to encode the orientation and other spatial features of each part. In the U-Net feature space, each part is either present or absent. Since all parts are present on this adversarial image, the U-Net can be fooled to detect the entire brain (H).

We can indeed use data augmentation to train U-Nets to detect objects with changed spatial features. We can also use adversarial training to prevent U-Nets from detecting adversarial images. But these inefficiencies lead to the need for a larger U-Net model. On the other hand, CapsNets handle the changed spatial features in a smarter way. This allows CapsNets, which are one order of magnitude smaller compared to U-Nets, to achieve similar results.^4^

## Appendix 4: Decision Choices to Develop Capsule Networks

To develop 3D CapsNets and make them work for volumetric brain MRI segmentation, we explored multiple architecture options, hyperparameters, loss functions, and implementation details to find optimal solutions. We used the validation set to explore these questions and select the best-performing model. We tested the best-performing model on the test set only once. Here are the design options that we explored and how we chose the winning options:

1. Network architecture: we built on the previous work by LaLonde et all^4^ to develop a 3D capsule network architecture. While Figure 1.A in the paper shows the architecture that performed best on the validation set, we explored the following options to get to the final architecture:
  a. Kernel size: we explored 3×3×3 and 5×5×5 kernels; the latter performed better.
  b. Downsampling method: we explored max-pooling versus 5×5×5 convolutions with stride of 2; the latter performed better.
  c. Upsampling method: we explored tri-linear interpolation versus 4×4×4 transposed convolutions with stride of 2; the latter performed better.
  d. Number of dynamic routing iterations to find agreeing pose vectors: we explored 1, 2, and 3 iternations for dynamic routing between capsule layers. As shown in Figure 1.A in the paper, the best-performing network uses 1 iteration for the first capsule layer, and 3 iterations for subsequent capsule layers.
  e. Number of capsule types in each layer: we tested up to 8 capsule types in the deepest part of the network, which performed best. Memory limitations did not let us increase the number of capsule types any further.
  f. Number of pose vector elements in each capsule: we tested up to 64 pose vector elements in the deepest part of the network, which performed best. Memory limitations did not allow us to further increase the number of pose vector elements.
2. Optimizer hyperparameters: we used Adam optimizer with dynamic scheduling of the learning rate. We explored the following ranges of hyperparameters to select the best-performing model over the validation set. Since the best-performing hyperparameters are detailed in Appendix 7, we are not listing them here again:
  a. Initial learning rate: we explored values ranging from 0.02 down to 0.001.
  b. Minimal learning rate: we explored values ranging from 0.001 down to 0.0001.
  c. Dynamic learning rate scheduling patience: if the model performance does not improve over a number of training epochs (in our case, mini-epochs), the learning rate scheduler decreases the learning rate by a factor. The number of epochs that the learning rate scheduler “waits” before decreases the learning rate, tolerating no improvement in performance over the validation set, is determined by the learning rate scheduler’s *patience*. We explored patience values ranging from 5 to 10.
  d. Learning rate decrement factor: when the model performance does not improve (over the validation set) after a number of epochs (determined by patience), the learning rate scheduler divides the learning rate by a number. We explored values ranging from 2 to 10.
3. Loss functions: we explored the following loss functions and found out that Dice loss perform best for our task:
  a. Dice loss
  b. Weighted average of Dice loss and cross-entropy loss
  c. Intersection over union loss
  d. Weighted average of intersections over union loss and cross-entropy loss
  e. Soft dice loss Our implementation of these loss functions is available at: https://github.com/Aneja-Lab-Yale/Aneja-Lab-Public-CapsNet/blob/main/loss_functions.py
4. Other design options:
  a. Batch size: we explored batch sizes ranging from 2 to 8. The batch size of 4 performed best over the validartion set.
  b. Patch size: we explored patch sizes ranging from 32×32×32 up to 128×128×128. While the larger patch size of 128×128×128 allows the network to use more contextual information in the image to segment each structure, larger patch sizes use more computational memory, forcing us to change other memory-intensive design options in the networks such as decreasing the number of capsule types in each layer or decreasing the number of pose elements in each capsule. Our experiments showed that a patch size of 64×64×64 led to the best-performing model over the validation set.
  c. Converting the activations in the final capsule layer into segmentations: we introduced a *forgiving paradigm* to convert final-layer activations into segmentations. This forgiving paradigm accelerated CapsNet training and made the training process stable. Appendix 5 details this paradigm.

## Appendix 5: Converting Final Layer Activations into Segmentations

The final layer of the 3D CapsNet is composed of one capsule channel that learns to activate capsules within the segmentation target and deactivate them outside the target. Activation of a capsule is determined by the length of its pose vector, which is a number between 0 and 1. The ground truth segmentations are coded similarly: pixels outside and inside the segmentation target are respectively coded by 0 and 1.

During testing, the length of the final layer’s pose vectors is thresholded at *T*:

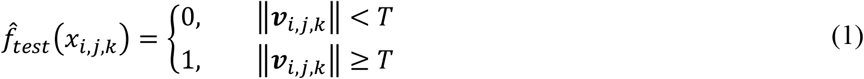

where 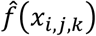 is the prediction of the CapsNet for the input voxel *x*_*i,j,k*_ and ‖***v***_*i,j,k*_‖ is the length of the final layer’s pose vector ***v***_*i,j,k*_ at the location (*i,j,k*) of the MRI volume (please note that ***v***_*i,j,k*_ is itself a function of *x*_*i,j,k*_, the function being the entire CapsNet that takes *x*_*i,j,k*_ as the input and gives ***v***_*i,j,k*_ as the output).

During training, the length of the final layer’s pose vector and each location (*i,j,k*) undergo a piecewise linear transform as follows:

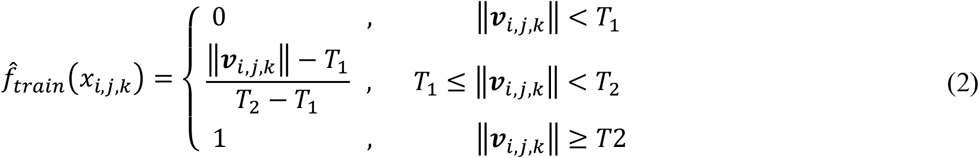

If we set *T* = 0.5 *T*_1_ = 0.1 and *T*_2_ = 0.9, we get the following diagrams for 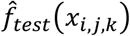 and 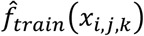 as functions of ‖***v***_*i,j,k*_‖:

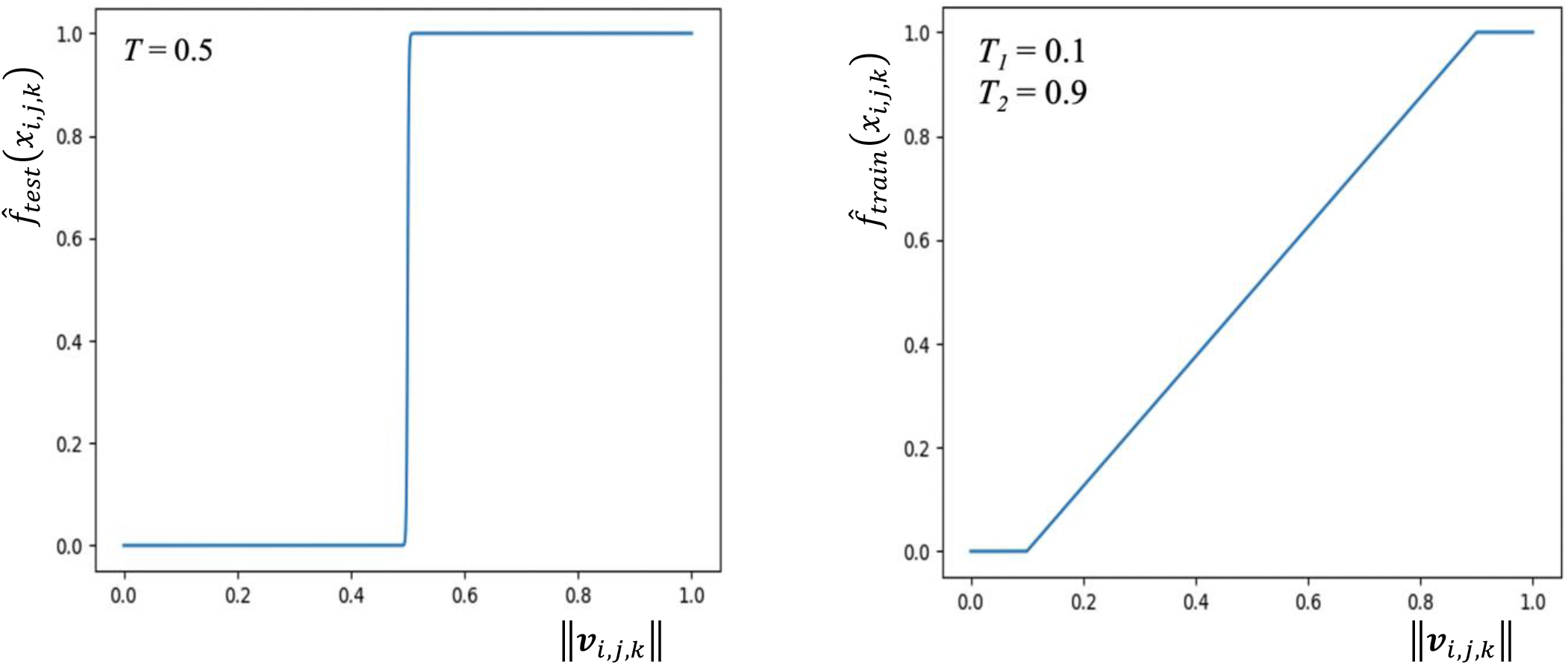

During training, the piecewise conversion (formula 2) enables a *forgiving paradigm* for the length of the final layer’s pose vectors: if the length of the vector is more than 0.9 for a voxel inside the segmentation target, the loss for that voxel would be zero. Intuitively, a pose vector with a length more than 0.9 for a voxel inside the segmentation target is considered “good enough”, so the training algorithm should not try to perfect the length of this vector to 1. Similarly, a pose vector with a length less than 0.1 is considered good enough for a voxel outside the segmentation target, so the training algorithm should not try to perfect the length of this vector to 0. This forgiving training paradigm makes the training of CapsNet stable because this paradigm does not try to perfect the length of the pose vectors of the final layer to 0’s and 1’s. In contrast, if a training paradigm tries to perfect the length of the pose vectors to 0’s and 1’s, that training paradigm becomes unstable because the pose vectors can assume a length close to 0 or 1, but not exactly 0 or 1. Remember that the pose vectors are generated by the *squash function*,^5^ which cannot generate vectors with a length equal to 0 or 1:

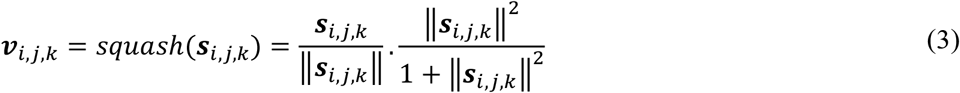

where ***s***_*i,j,k*_ is the total input to the final layer capsule at the location (*i,j,k*), and ***v***_*i,j,k*_ is the pose vector of the final layer capsule at that location.

Our experiments show that training with the forgiving paradigm is more stable and leads to faster convergence. When we did not convert the length of the pose vector ‖***v***_*i,j,k*_‖ using the conversion function (formula 2), CapsNet training became unstable. Here we show the evolution of the training set and the validation set losses during 10 epochs of training, with and without the forgiving paradigm:

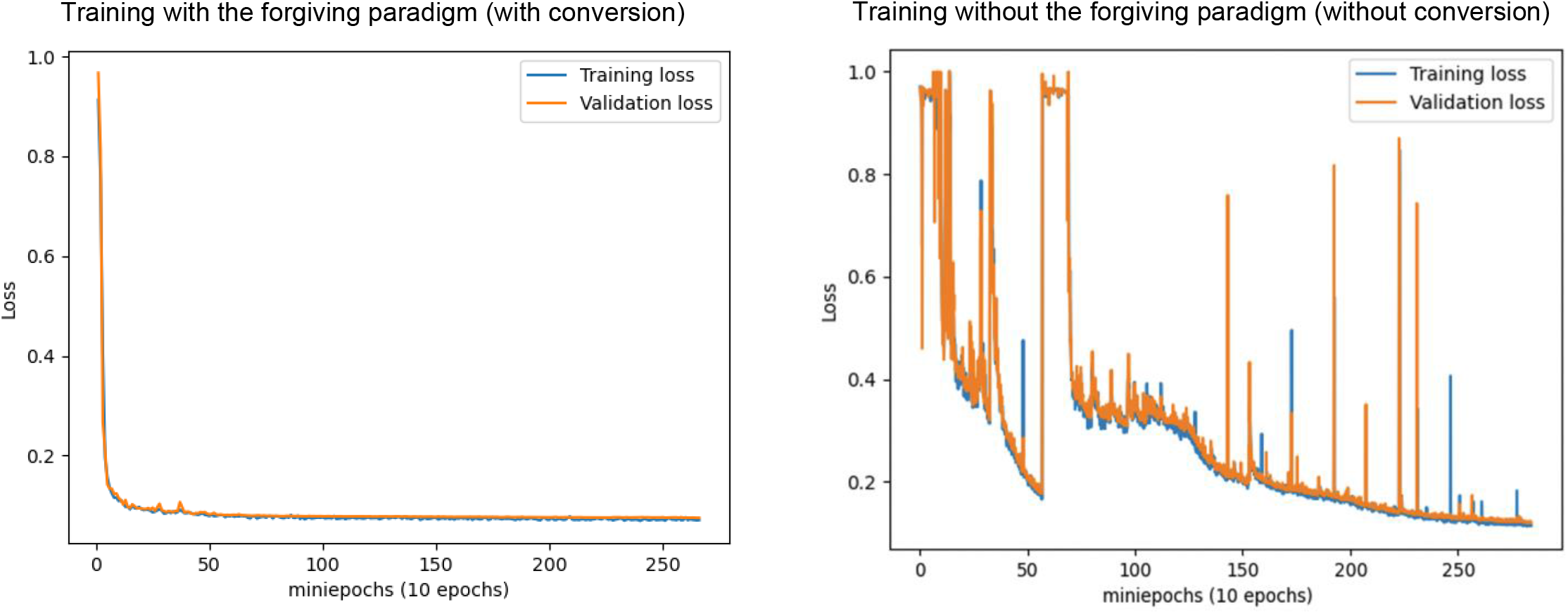

We additionally searched for the optimal conversion functions. The piecewise linear function led to the most stable training and fastest convergence. Here we describe other functions that we studied (together with their plots) so that other groups would be aware of these conversion functions that we think are suboptimal for this task:

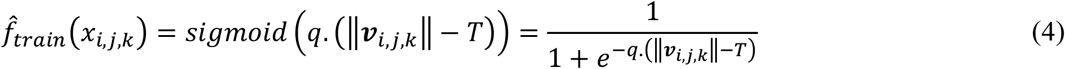

We set *T* = 0.5 and tried different values for *q* (10, 15, and 20):

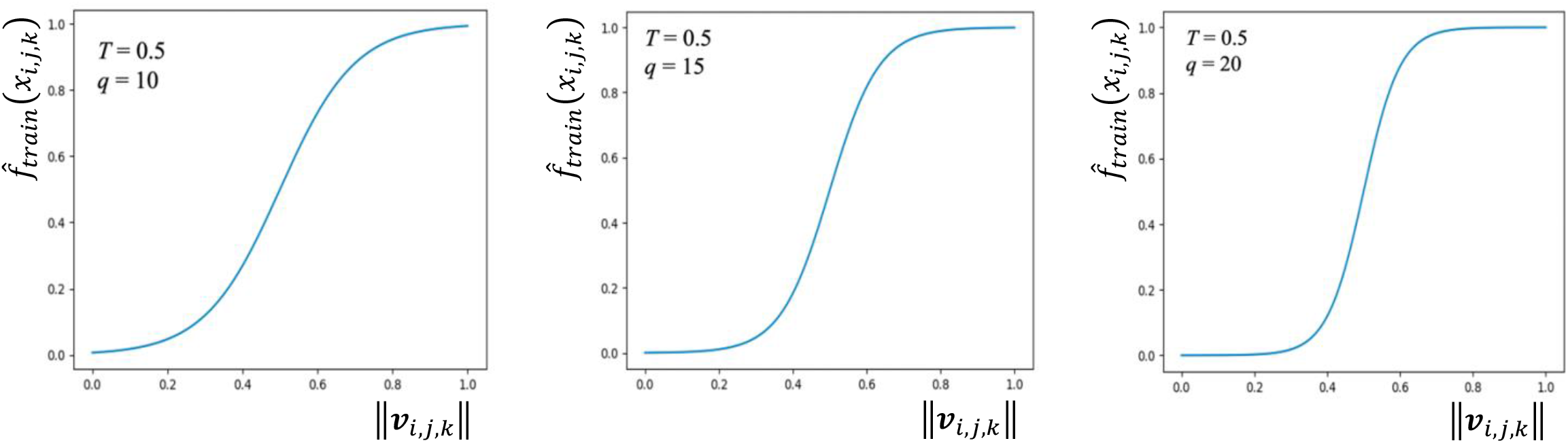

We also examined the piecewise conversion function (formula 2) with values for *T*_1_ and *T*_2_ other than 0.1 and 0.9:

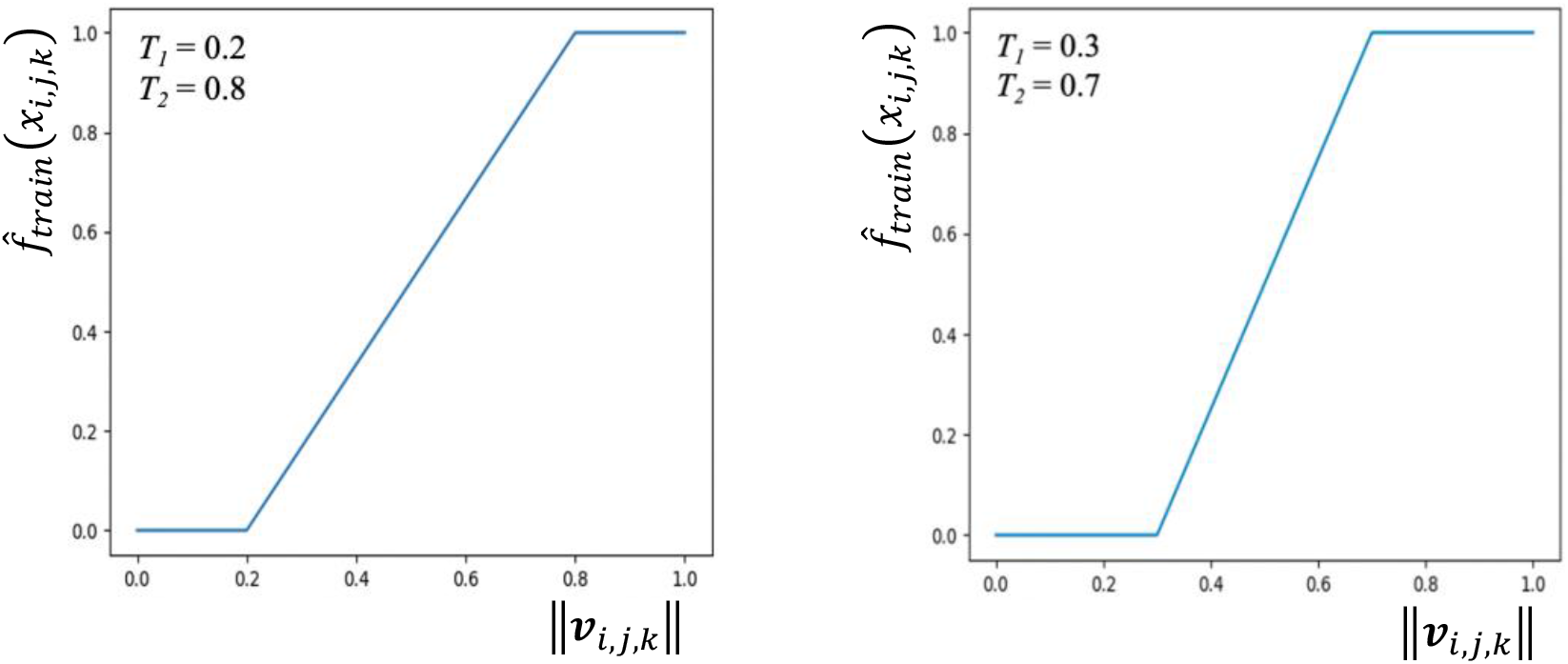

None of these conversion functions was as effective as the piecewise function with *T*_1_ = 0.1 and *T*_2_ = 0.9 in improving the stability and convergence of CapsNet training.

## Appendix 6: Findings Agreeing Pose Vectors

Let’s assume the previous capsule layers has six capsule channels, each outputting the vote vector of a part (*v*_*1*_ to *v*_*6*_). To find the vote vectors that agree, we first compute the vector summation of all vote vectors (*v*):

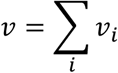

Then, we compute the inner products between each vote vector *v*_*i*_ and the sum *v*, yielding weights for each vote vector *w*_*i*_:

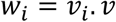

*w*_*i*_ = *v*_*i*_. *v* Please note that each *w*_*i*_ is a scalar. Next, we re-compute the vector sum *v* using the weighted average of the vote vectors using weights *w*_*i*_ computed in the previous step :

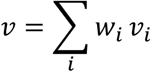

This process is often repeated for three iterations. The number of iterations is a hyperparameter that should be set between capsule layers. This whole process increases the weights of the vectors that align with the sum (*v*_*1*_, *v*_*2*_, and *v*_*6*_ in this example) and decreases the weights of the vectors that do not align with the sum (*v*_*3*_, *v*_*4*_, and *v*_*5*_ in this example).

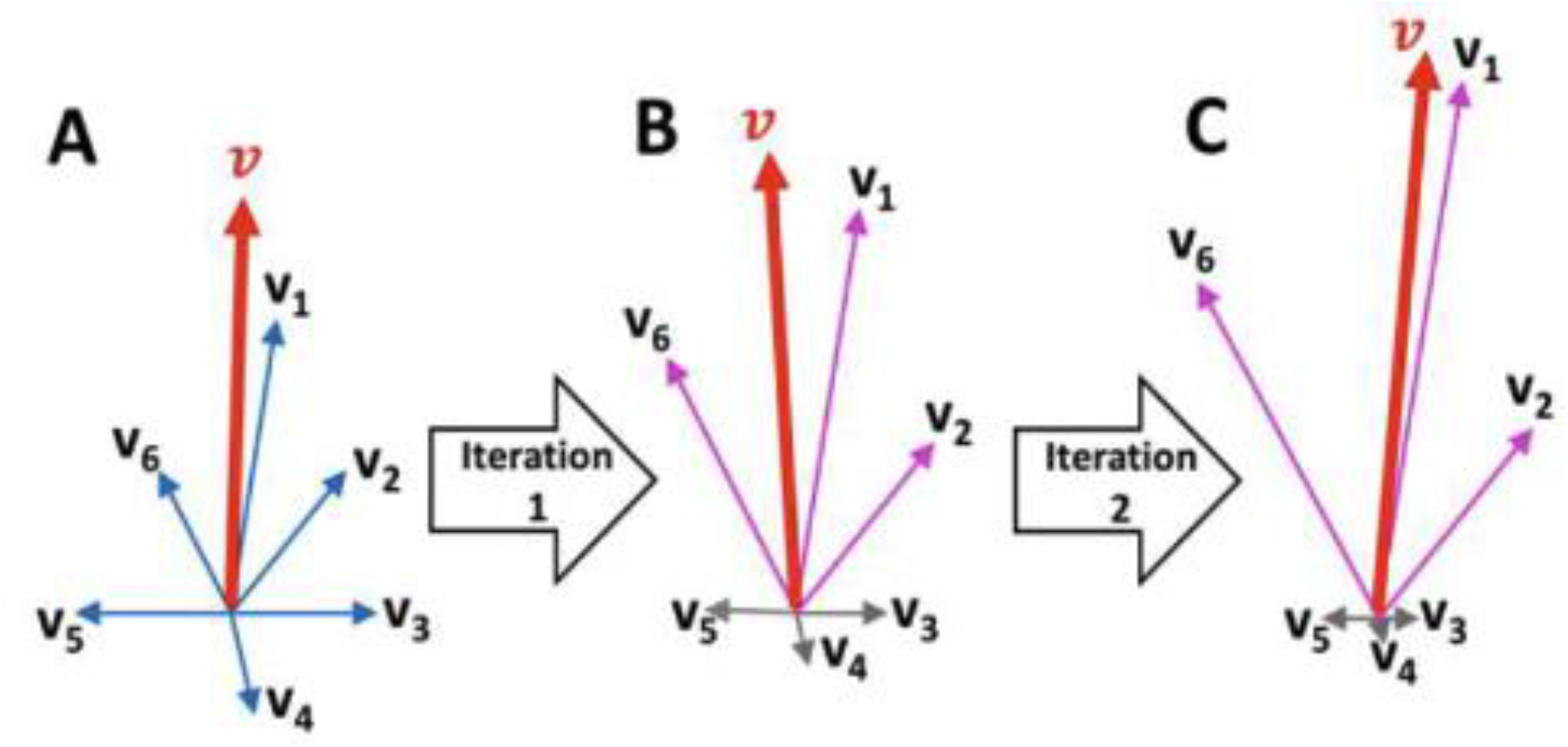

## Appendix 7: Training hyperparameters

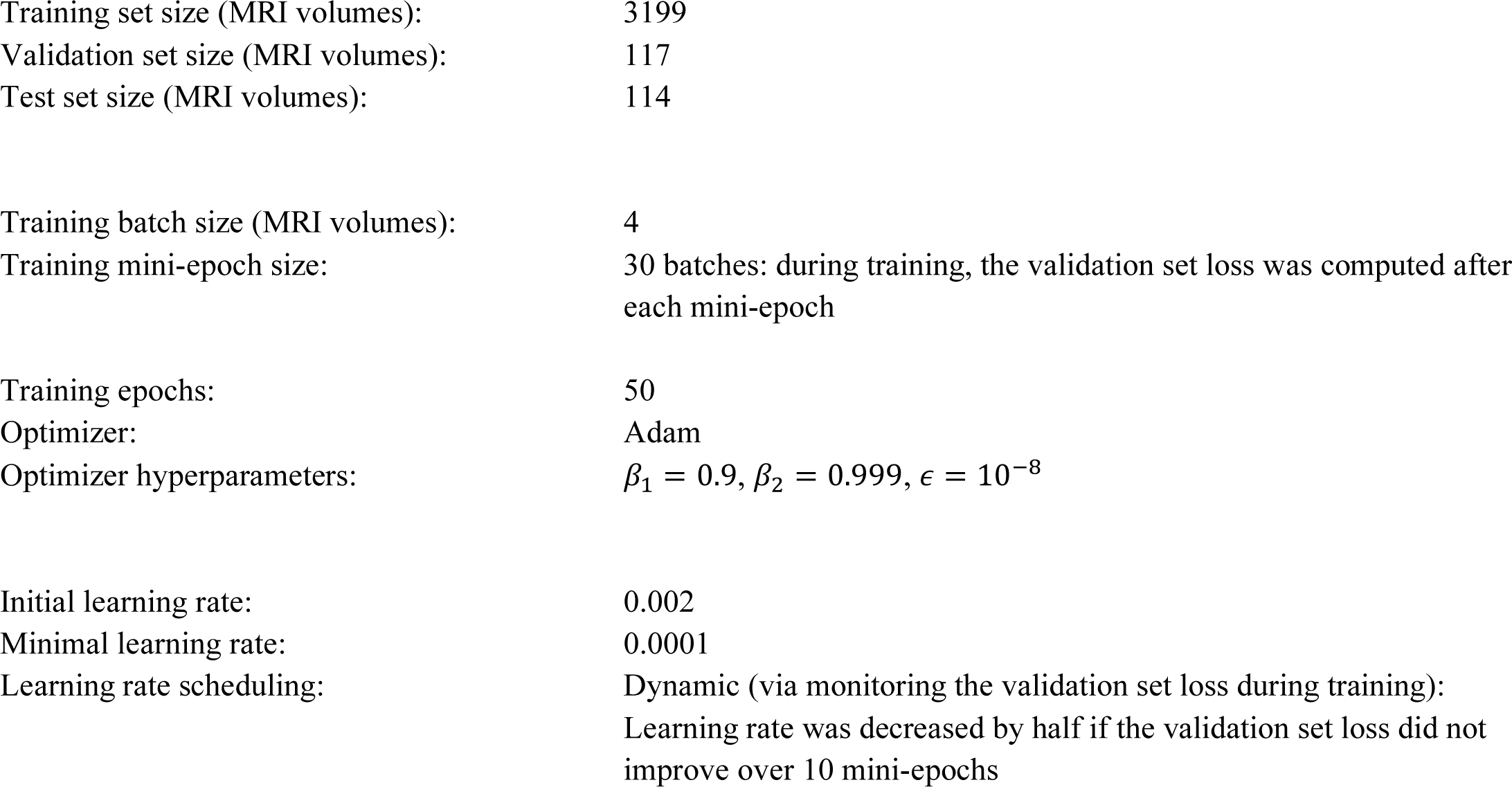

|  |  |
| --- | --- |
| Training set size (MRI volumes): | 3199 |
| Validation set size (MRI volumes): | 117 |
| Test set size (MRI volumes): | 114 |
| Training batch size (MRI volumes): | 4 |
| Training mini-epoch size: | 30 batches: during training, the validation set loss was computed after each mini-epoch |
| Training epochs: | 50 |
| Optimizer: | Adam |
| Optimizer hyperparameters: | $\beta_1 = 0.9, \beta_2 = 0.999, \epsilon = 10^{-8}$ |
| Initial learning rate: | 0.002 |
| Minimal learning rate: | 0.0001 |
| Learning rate scheduling: | Dynamic (via monitoring the validation set loss during training):<br>Learning rate was decreased by half if the validation set loss did not improve over 10 mini-epochs |

